# Triglyceride and Glucose Index and Sex Differences in Relation to Cognitive Impairment in Hypertensive Patients without Diabetes

**DOI:** 10.1101/2023.11.06.23298189

**Authors:** Rufei Liu, Wenli Cheng

**Author notes:** **Correspondence:** Wenli Cheng.

## Abstract

**Background:** Previous observational studies in patients with diabetes have examined the relationship between insulin resistance (IR) and cognitive outcomes and failed to find that IR is associated with cognitive function. And the triglyceride-glucose (TyG) index, which measured serum triglyceride (TG) and fasting blood glucose (FBG), has been suggested as a surrogate reliable marker of IR and widely used due to its convenience and cost-effectiveness.However, to the best of our knowledge, the relationship between IR and intensive blood pressure control in specific cognitive outcomes has never been investigated.

**Methods:** To fill this knowledge gap, we analyzed the relationship between TYG levels and cognitive outcomes in hypertensive patients within the Systolic Blood Pressure Intervention Trial (SPRINT). The SPRINT evaluated the impact of intensive blood pressure control (systolic blood pressure < 120 mmHg) versus standard blood pressure control (systolic blood pressure < 140 mmHg). The Cox proportional risk regression was used to investigate the association between different TYG status and clinical outcomes. Additional stratified analyzes were performed to evaluate the robustness of gender difference.

**Results:** A total of 9,323 participants (6016 [64.53%] males and 3307 [35.47%] females) with hypertension from the SPRINT research were included in the analysis. The median follow-up period was 3.26 years. Our population was divided into three groups according to the size of the TyG index. The low TyG group was the reference. Sensitivity analyzes showed that in the SPRINT, the TyG index was significantly associated with the risk of cognitive outcomes across various subgroups. There was no significant interaction in the confounders.

**Conclusions:** In this cohort study, results suggest that patients with TyG levels higher had lower risk of probable dementia, but this study tested association, not causation. Our results demonstrated that in patients with hypertension, the association between TyG and risk of probable dementia is L-shaped.

## INTRODUCTION

Cognitive decline is a preclinical stage of dementia, which is characterised by deterioration in memory, comprehension, attention and calculation[1, 2]. The burden of cognitive decline on the global public health system is enormous. In China, the prevalence of dementia and mild cognitive impairment (MCI) was 6.04% and 15.54% respectively from 2015 to 2018[2]. Currently, identifying risk factors to prevent cognitive decline at an early stage is essential, as there are no effective drugs for dementia[2].

Growing evidence links insulin resistance (IR) with risk of cognitive decline[3], the mechanisms that underlie the decline in cognitive function have also been elucidated by clinical trials.[4]. Nevertheless, most previous studies have focused on the association between IR and cognitive impairment and dementia[2, 3], not cognitive decline directly. In addition, the epidemiological evidence in this area is still limited. It is noteworthy that previous research considered insulin clamp techniques and intravenous glucose tolerance tests to be the standards for the diagnosis of IR[5-7], although in reality these were not routine procedures. Therefore, the triglyceride-glucose (TyG) index, which measured serum triglyceride (TG) and fasting blood glucose (FBG)[8], has been suggested as a surrogate reliable marker of IR[5, 6] and widely used due to its convenience and cost-effectiveness[3]. Although as a surrogate indicator of IR, TyG index can detect the relationship of IR and cardiovascular diseases[9] or certain types of cancer, which has been observed by using the gold standards of IR. However, in assessing the relationship between insulin resistance and cognitive decline, the reliability of the TyG index as a surrogate marker for insulin resistance remains uncertain.

The population included in the previous studies is the general population[10]. Older people, who usually have a higher prevalence of cardiovascular risk factors, have rarely been included in trials. Moreover, extant evidence suggests the presence of gender disparities in insulin resistance and cognitive decline. Women were more likely to suffer from IR[11, 12] and cognitive decline[2] than men. It is unclear whether IR is associated with cognitive decline according to sex. To better study these issues, we used the data from the Systolic Blood Pressure Intervention Trial (SPRINT)[13] to evaluate the relationship between TyG index and cognitive impairment in a hypertension population and further explore the correlation between gender differences.

## METHOD

We performed a post-hoc analysis of the SPRINT. The limited dataset was obtained from the National Institutes of Health Biologic Specimen and Data Repository Information Coordinating Center (https://biolincc.nhlbi.nih.gov/studies/sprint/).

### Study Population

SPRINT was a randomized, controlled trial conducted at 102 clinical sites in the United States. The rationale, design, and main results of SPRINT have been previously published[14]. Briefly, SPRINT was designed to test whether the intensive management of systolic BP to < 120 mmHg reduces cardiovascular disease events compared with standard BP management (< 140 mmHg). The recruited participants were between the ages of 50 and 75 and had at least one of the following: presence of clinical or subclinical cardiovascular disease other than stroke; chronic kidney disease (defined as eGFR 20–59 ml/min/1.73 m2); Framingham risk score for 10-year CVD risk ≥15% based on laboratory work done in the last 12 months; or if patients were aged 75 years or older. Because the blood pressure trial of ACCORD study did not come to a good conclusion within the diabetic patients[15]. Exclusion criteria were that patients had type 2 diabetes, prior stroke, and standing systolic BP <110 mmHg at the screening visit. The SPRINT showed that intensive blood pressure management significantly reduced cardiovascular mortality and all-cause mortality compared to standard management.

### Exposure Variables

TyG index was defined as TyG=Ln [fasting triglycerides (mg/dl) ×fasting glucose (mg/dl)/2]. We used the baseline fasting triglycerides and fasting glucose to calculate the TyG index[16, 17]. We divided the population into three groups according to the size of the TyG index. The first group was the reference. The primary cognitive outcome was the occurrence of probable dementia. The secondary cognitive outcomes were the occurrence of MCI and a composite of probable dementia or MCI. Cognitive status of each participant was assessed at baseline and during follow-up by series of tests including Montreal Cognitive Assessment, Logical Memory forms I and II subtests of the Wechsler Memory Scale, Digit Symbol Coding Test of the Wechsler Adult Intelligence Scale, and Functional Activities Questionnaire. Participants were diagnosed as no cognitive impairment, MCI, or probable dementia according to standardized criteria by two adjudicators.[18, 19] More details have been published elsewhere.[20] The outcomes were adjudicated.

### Statistical Analysis

For categorical variables, baseline patient characteristics and outcomes were expressed as frequencies and percentages. For continuous variables, mean and standard deviation (SD) or median and interquartile range (IQR) were used, depending on the normal distribution of the data (evaluated using normal Q-Q charts). To compare categorical variables, we used chi-squared analysis. For comparison of continuous variables, we used analysis of variance or Mann-Whitney U test, depending on the distribution type.

The variables that were adjusted for in this study were selected on the basis of their clinical importance. To estimate the association of baseline TyG index with the risk of primary and secondary cognitive outcomes, hazard ratios (HRs) and 95% confidence intervals (CIs) were calculated using three multivariable Cox proportional hazards regressions. Scaled Schoenfeld residuals were used to test the validity of the proportionality assumption. Model 1 was an unadjusted model. Model 2 was a model with adjustment for age, treatment arm and ethnicity. Model 3 was further adjusted for age, treatment arm, ethnicity, baseline body mass index, smoking status, chronic kidney disease (CKD) subgroup, cardiovascular disease (CVD) subgroup, number of antihypertensive agents, aspirin used, and statin used at baseline. To account for the TyG index as a continuous variable, we constructed a Cox proportional hazards regression model (Model 3). The TyG index was used to calculate the HR for outcomes. Then restricted cubic spline models were built to detect any non-linear relationship between TyG index and probable dementia. We used two-piecewise linear regression models to elucidate how the associations differed by the threshold point. The threshold value was estimated by trying all possible values and choosing the threshold point with the highest likelihood. A logarithmic likelihood ratio test was employed to compare the differences in associations when using one-line linear regression models vs. two-piecewise linear regression models. We performed the interaction and stratified analyses by sex, treatment arm, age (< 75 years and ≥ 75 years), systolic blood pressure tertile (≤ 132 mmHg, 132– 145 mmHg, and ≥ 145 mmHg), Framingham 10-y cardiovascular disease risk score (≤ 15%, > 15%), smoking status, CVD subgroup, CKD subgroup, TIA subgroup, Black race, aspirin use, statin use and number of distinct anti-hypertensive agents.

All analyses were performed using statistical software packages R (The R Foundation; http://www.R-project.org) and EmpowerStats (X&Y Solutions, Inc., Boston, MA, USA; http://www.empowerstats.com). P values < 0.05 (two-sided) were considered statistically significant.

## RESULTS

### Baseline Characteristics of Included Hypertension Patients

There are 38 patients for whom the TyG index cannot be calculated out of a total of 9,323 patients with hypertension in the SPRINT study. The median follow-up was 3.26 years. The mean age for all the participants was 67.92 ± 9.42 years; 3,307 (35.47%) participants were females, 2,785 (29.87%) were black, 1,869 (20.05%) had a history of CVD, and 2,645 (28.37%) had a history of CKD. Individuals with a low TyG score constituted 33.29% (3104 of 9,323) of this cohort and were likely to be older (mean [SD] age, 69.15 [9.25]), female (1174 [37.82%]), with a higher prevalence of cardiovascular risk factors (mean [SD] systolic blood pressure (140.13 [15.63] mmHg); mean [SD] heart rate (65.17 [11.27] bpm)) compared with those with higher TyG scores. High TyG group patients had a higher Framingham 10-y cardiovascular disease risk score. Individuals with a low TyG score constituted 33.29% (3104 of 9,323) of this cohort also had lower median (IQR) triglyceride levels (69.00 [54.00-89.0] mg/dL; to convert to millimoles per liter, multiply by 0.0113) and lower mean (SD) LDL-C levels (120.24 [37.21] mg/dL; to convert to millimoles per liter, multiply by 0.0259) **(Table 1)** than those with higher TyG scores. The between-group differences were consistent across the components of the primary outcome and other prespecified secondary outcomes. **(Table 1)** After follow-up, 155 (4.99%) primary outcome occurred in the low TyG group, 106(3.73%) in the middle group, and 87 (3.04%) in the high TyG group. Decrease in primary outcome observed as TyG score increases (P=0.011). **Table 1** provides the detailed baseline characteristics of the patients with hypertension included in the study population.

**Table 1.**
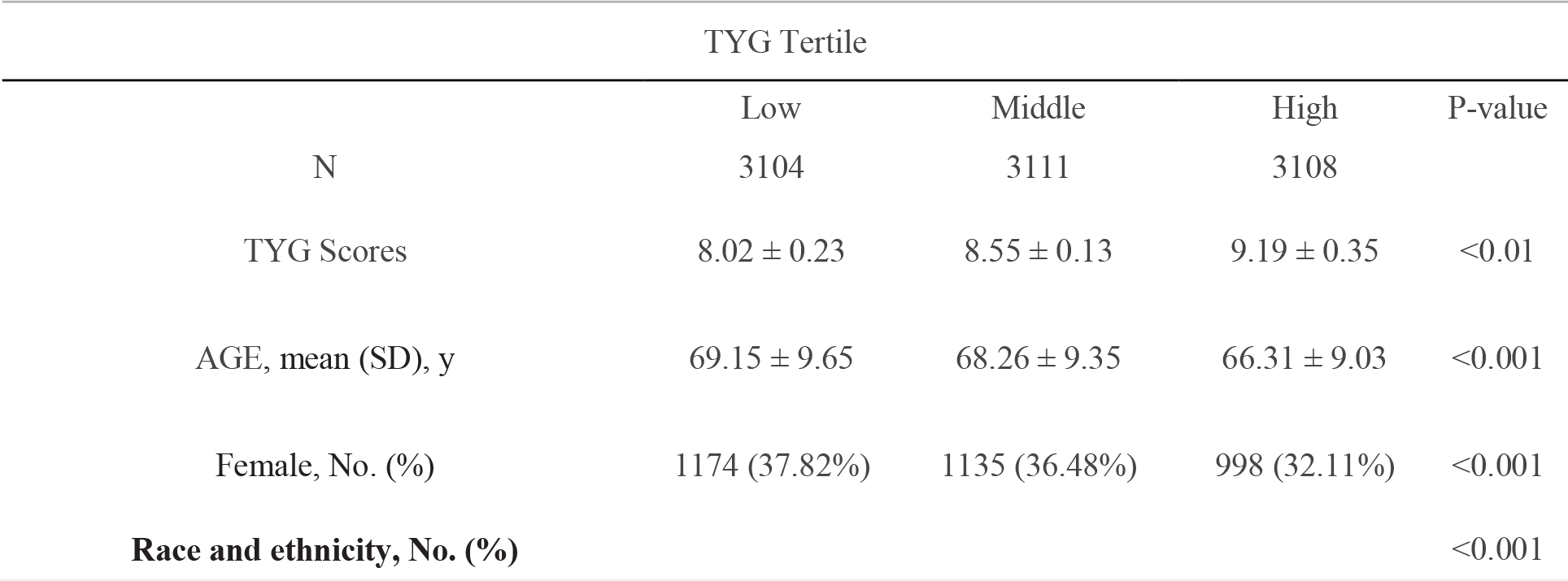
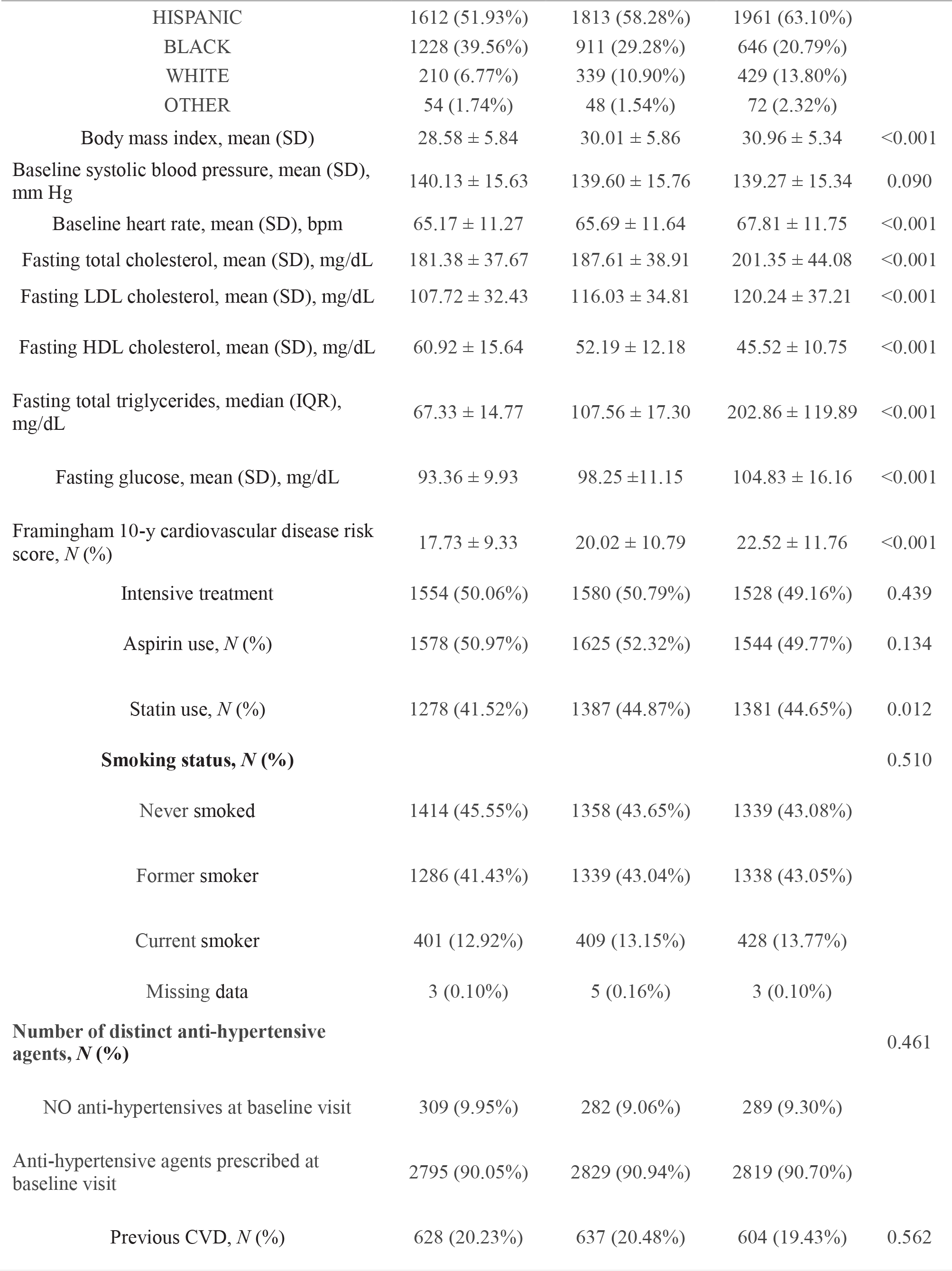
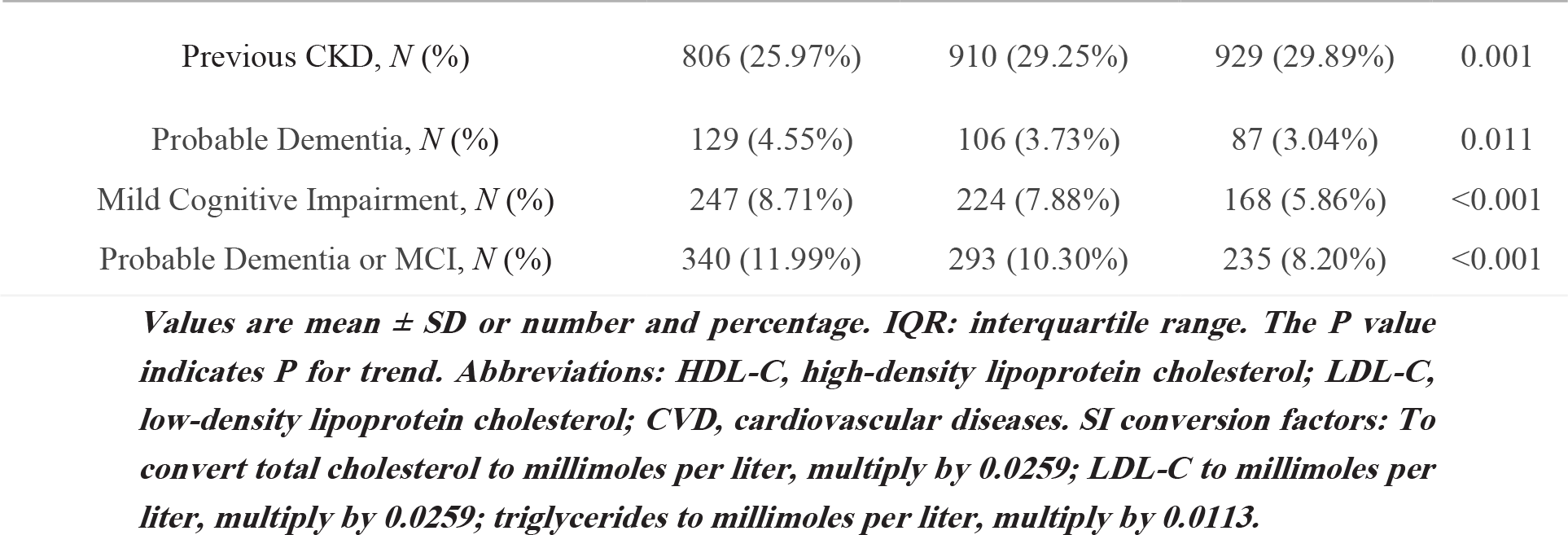
Baseline characteristics and crude end points of the study participants.

|  | TYG Tertile |  |  | P-value |
| --- | --- | --- | --- | --- |
|  | Low | Middle | High |  |
| N | 3104 | 3111 | 3108 |  |
| TYG Scores | 8.02 ± 0.23 | 8.55 ± 0.13 | 9.19 ± 0.35 | <0.01 |
| AGE, mean (SD), y | 69.15 ± 9.65 | 68.26 ± 9.35 | 66.31 ± 9.03 | <0.001 |
| Female, No. (%) | 1174 (37.82%) | 1135 (36.48%) | 998 (32.11%) | <0.001 |
| **Race and ethnicity, No. (%)** |  |  |  | **<0.001** |
| HISPANIC | 1612 (51.93%) | 1813 (58.28%) | 1961 (63.10%) |  |
| BLACK | 1228 (39.56%) | 911 (29.28%) | 646 (20.79%) |  |
| WHITE | 210 (6.77%) | 339 (10.90%) | 429 (13.80%) |  |
| OTHER | 54 (1.74%) | 48 (1.54%) | 72 (2.32%) |  |
| Body mass index, mean (SD) | 28.58 ± 5.84 | 30.01 ± 5.86 | 30.96 ± 5.34 | <0.001 |
| Baseline systolic blood pressure, mean (SD), mm Hg | 140.13 ± 15.63 | 139.60 ± 15.76 | 139.27 ± 15.34 | 0.090 |
| Baseline heart rate, mean (SD), bpm | 65.17 ± 11.27 | 65.69 ± 11.64 | 67.81 ± 11.75 | <0.001 |
| Fasting total cholesterol, mean (SD), mg/dL | 181.38 ± 37.67 | 187.61 ± 38.91 | 201.35 ± 44.08 | <0.001 |
| Fasting LDL cholesterol, mean (SD), mg/dL | 107.72 ± 32.43 | 116.03 ± 34.81 | 120.24 ± 37.21 | <0.001 |
| Fasting HDL cholesterol, mean (SD), mg/dL | 60.92 ± 15.64 | 52.19 ± 12.18 | 45.52 ± 10.75 | <0.001 |
| Fasting total triglycerides, median (IQR), mg/dL | 67.33 ± 14.77 | 107.56 ± 17.30 | 202.86 ± 119.89 | <0.001 |
| Fasting glucose, mean (SD), mg/dL | 93.36 ± 9.93 | 98.25 ± 11.15 | 104.83 ± 16.16 | <0.001 |
| Framingham 10-y cardiovascular disease risk score, *N* (%) | 17.73 ± 9.33 | 20.02 ± 10.79 | 22.52 ± 11.76 | <0.001 |
| Intensive treatment | 1554 (50.06%) | 1580 (50.79%) | 1528 (49.16%) | 0.439 |
| Aspirin use, *N* (%) | 1578 (50.97%) | 1625 (52.32%) | 1544 (49.77%) | 0.134 |
| Statin use, *N* (%) | 1278 (41.52%) | 1387 (44.87%) | 1381 (44.65%) | 0.012 |
| **Smoking status, *N* (%)** |  |  |  | 0.510 |
| Never smoked | 1414 (45.55%) | 1358 (43.65%) | 1339 (43.08%) |  |
| Former smoker | 1286 (41.43%) | 1339 (43.04%) | 1338 (43.05%) |  |
| Current smoker | 401 (12.92%) | 409 (13.15%) | 428 (13.77%) |  |
| Missing data | 3 (0.10%) | 5 (0.16%) | 3 (0.10%) |  |
| **Number of distinct anti-hypertensive agents, *N* (%)** |  |  |  | 0.461 |
| NO anti-hypertensives at baseline visit | 309 (9.95%) | 282 (9.06%) | 289 (9.30%) |  |
| Anti-hypertensive agents prescribed at baseline visit | 2795 (90.05%) | 2829 (90.94%) | 2819 (90.70%) |  |
| Previous CVD, *N* (%) | 628 (20.23%) | 637 (20.48%) | 604 (19.43%) | 0.562 |
| Previous CKD, *N* (%) | 806 (25.97%) | 910 (29.25%) | 929 (29.89%) | 0.001 |
| Probable Dementia, *N* (%) | 129 (4.55%) | 106 (3.73%) | 87 (3.04%) | 0.011 |
| Mild Cognitive Impairment, *N* (%) | 247 (8.71%) | 224 (7.88%) | 168 (5.86%) | <0.001 |
| Probable Dementia or MCI, *N* (%) | 340 (11.99%) | 293 (10.30%) | 235 (8.20%) | <0.001 |

*Values are mean ± SD or number and percentage. IQR: interquartile range. The P value indicates P for trend. Abbreviations: HDL-C, high-density lipoprotein cholesterol; LDL-C, low-density lipoprotein cholesterol; CVD, cardiovascular diseases. SI conversion factors: To convert total cholesterol to millimoles per liter, multiply by 0.0259; LDL-C to millimoles per liter, multiply by 0.0259; triglycerides to millimoles per liter, multiply by 0.0113.*

### Tertiles of TyG Index and Cognitive Outcomes

The association between the TyG index and cognitive outcomes in patients with hypertension is presented in Table 2. In Model 3, the third tertile has the lowest risk of probable dementia (HR, 0.71; 95% CI, 0.51–0.99; P=0.0405). When the TyG index grouping was regarded as a continuous variable, this trend did not change, although the statistical difference was not significant.

**Table 2.**
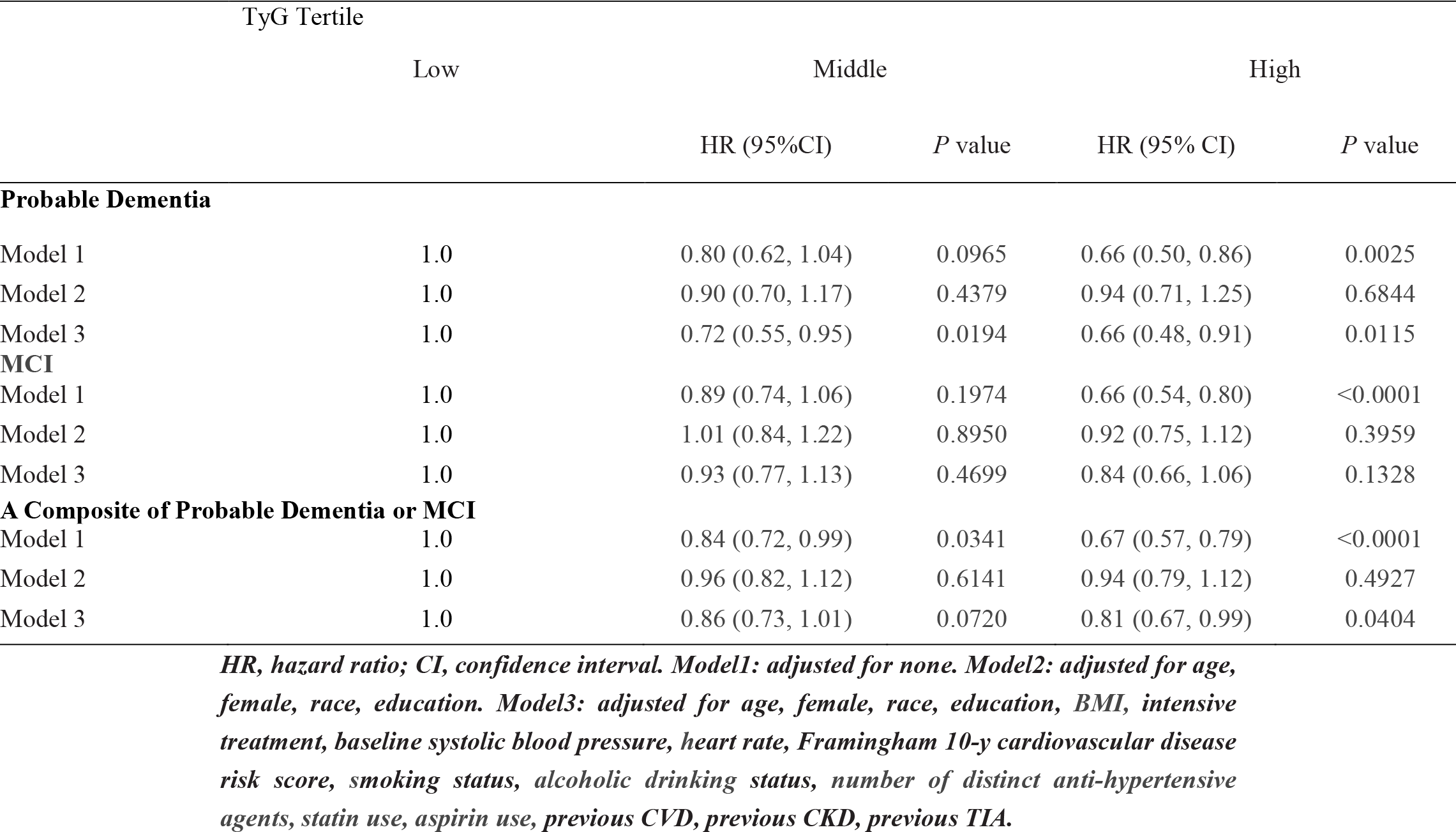
Association Between TyG and Cognitive Outcomes.

### TyG Index as a Continuous Variable and Cognitive Outcomes

As shown in Table 3, when we used the TyG index as a continuous covariate, with increase in TyG index decreased the risk of probable dementia, although the statistical difference was not significant (HR, 0.70; 95% CI, 0.48–1.02; P=0.0665). Restricted cubic splines were used to flexibly model and visualize the relationship between the TyG index and probable dementia. With the increase of the TyG index, the risk of probable dementia decreased. When the TyG index was close to nine, the trend of increasing the risk of probable dementia slowed down (Figure 1). There was no interaction between the sex and TyG index (P for interaction= 0.6169). Next, we used the two-stage linear regression model to calculate the threshold effect. Table 3 shows the results of the two-stage linear regression model. The inflection point was 8.94 in all participants; on the left inflection point, the effect size, 95% CI, and P value were 0.65, 0.46–0.92, and=0.0148, respectively; on the right inflection point, HR, 1.34; 95% CI, 0.74–2.43; P=0.3394. However, the log likelihood ratio test was 0.072. This means that the two-stage linear regression model was not better than the one-line linear regression models.

**Table 3.** Results of two-piecewise linear-regression model.

|  | Male |  | Female |  | Total |  |
| --- | --- | --- | --- | --- | --- | --- |
| One linear-regression mode | 0.70 (0.48, 1.02) | 0.0665 | 0.99 (0.61, 1.59) | 0.9552 | 0.79 (0.60, 1.04) | 0.0975 |
| Inflection point (K) | 8.92 |  | 9.09 |  | 8.94 |  |
| <K Effect size b (95% CI) | 0.57 (0.36, 0.92) | 0.0197 | 0.79 (0.47, 1.34) | 0.3921 | 0.65 (0.46, 0.92) | 0.0148 |
| >K Effect size b (95% CI) | 1.14 (0.54, 2.37) | 0.7359 | 3.11 (0.90, 10.72) | 0.0727 | 1.34 (0.74, 2.43) | 0.3394 |
| Log likelihood ratio test | 0.166 |  | <0.001 |  | 0.072 |  |

*Two-piecewise linear-regression model was used to calculate the threshold effect of the TyG index. If the log likelihood ratio test >0.05, it means the two-piecewise linear regression model is not superior to the single-line linear regression model.*

**FIGURE 1.**
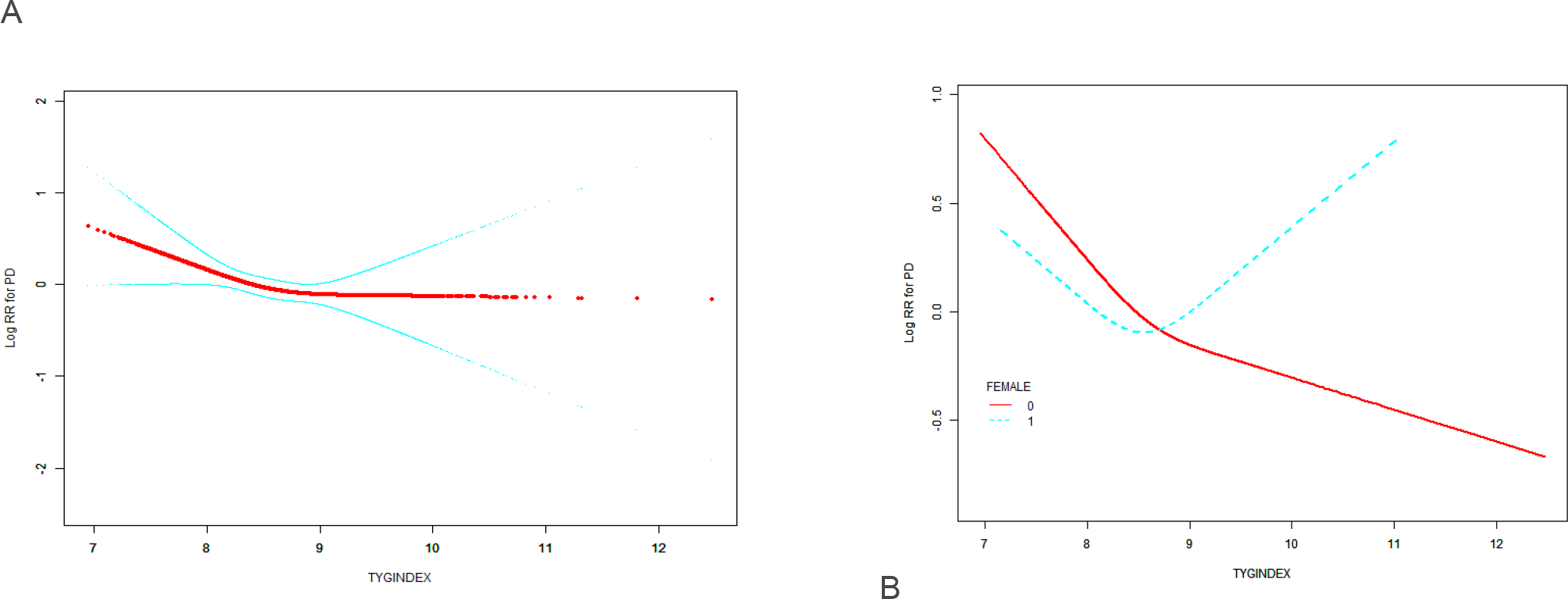
Relationship between TyG index and probable dementia. (A) Relationship between TyG index and probable dementia in all patients. The red line is the trend line and the blue line is the 95% confidence interval. (B) Relationship between TyG index and probable dementia grouped by sex. Male: red line; Female: blue line.

### Interaction and Sensitivity Analyses

The results of the interaction and stratified analyses are presented in Figure 2. Generally, the TyG index was significantly associated with the risk of cognitive outcomes across various subgroups. There was no significant interaction in the confounders.

**Figure 2:**
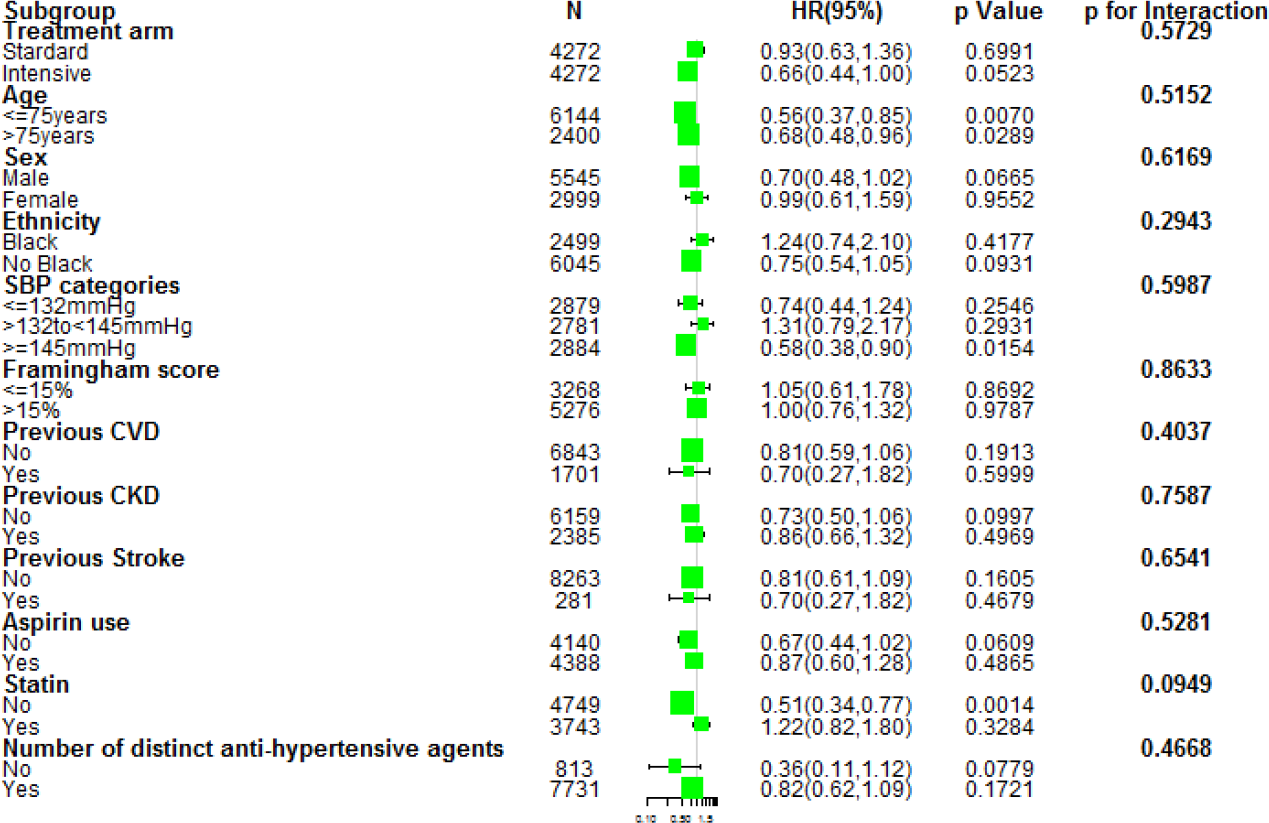
Subgroup analysis of the association between the different TYG levels and risk of probable dementia. Each subgroup analysis was adjusted for all factors in Model 3, except for the stratification factor itself.

## DISCUSSION

This study aimed to investigate how TyG index relates to cognitive impairment in people with high blood pressure without diabetes, and how it relates to sex. In this post-hoc analysis, we observed that a low baseline TyG index was associated with the risk of cognitive outcomes in elderly hypertensive patients without diabetes. And there was no significant interaction between sex and the TyG index.

Over the past few years, several observational studies have examined the relationship between IR and cognitive outcomes. Some studies have failed to find that IR is associated with cognitive function. For example, a similar study focusing on T2D patients and using HOMA2-IR to calculate the IR index came to a similar conclusion as ours, that HOMA2-IR was not associated with cognitive performance.[21, 22] However, our patients tended to explain the relationship between IR and cognitive dysfunction more simply, compared with people with diabetes. Previously, researchers evaluated the relationship between IR (using the homeostasis model assessment [HOMA] index) and cognitive decline among 3342 participants in the Rotterdam study sample: in participants without a history of diabetes, rising IR was not associated with cognitive function.[23] The TyG index calculated based on fasting triglycerides and blood glucose is a reliable alternative biomarker for evaluating insulin resistance. The accuracy of TyG index in diagnosing insulin resistance is consistent with Hyperinsulinemic-Euglycemic Clamp (HIEC) and Homeostatic Model Assessment of Insulin Resistance (HOMA-IR).[24] To the best of our knowledge, this study is the first study to evaluate the relationship between the TyG index and adverse outcomes within the population of hypertensive patients without diabetes. Furthermore, previous studies have shown that IR is the main mechanism of brain abnormalities in magnetic resonance imaging (MRI) of patients with or without diabetes.[25, 26] Neverthless, Ji Hee Yu, et al.[27] examined whether long-term hyperglycemia, insulin resistance, or secretory function is associated with brain atrophy and cognitive decline in subjects from the Ansan cohort of the Korean Genome Epidemiology Study (KoGES), finding time-weighted HOMA-IR level during a 10-year period did not show an association with brain atrophy, unlike baseline insulin resistance markers. Similarly, a longitudinal study reported that there was no correlation between insulin secretion assessed by insulinogenic index (IGI) obtained through oral glucose tolerance test (OGTT) and cognitive function of participants without diabetes.[24]

It can be explained from two aspects. On the one hand, in addition to insulin resistance, diabetes can also cause cognitive dysfunction through hyperglycemia, vascular diseases, hypoglycemia and other ways. Hence, a variety of pathogenesis greatly increase the possibility of diabetes as a risk factor. On the other hand, TYG inflects peripheral insulin resistance. It is easy to understand that the ability of peripheral insulin secretion may not represent the central insulin action or concentration. That is because (1) the transporters of the blood-brain barrier exhibit saturability, indicating that the correlation between insulin levels in the bloodstream and those in the central nervous system is non-linear, (2) the CNS is not part of the classical negative feedback loop that regulates insulin release, (3) the effect of CNS insulin on peripheral insulin and glucose levels is opposite to that of peripheral insulin. In fact, insulin resistance in the periphery and CNS occur simultaneously, is not always correct. The pioneering study by Talbot et al. showed that the presence of peripheral insulin resistance does not imply the presence of CNS insulin resistance[28]. This study shows that the majority of people with Alzheimer’s disease (AD) have a resistance to insulin receptors in their brains, whereas people with diabetes do not have such a resistance.This is an indication that there is CNS insulin resistance even in the absence of peripheral insulin resistance. CNS insulin resistance is an entity independent of peripheral blood insulin resistance. Although both can lead to cognitive decline and ultimately lead to AD, they are likely to be achieved through different pathways. Decrease in CSF insulin concentration in obese individuals and AD patients indicates a decrease in insulin transport through the blood-brain barrier. Peripheral insulin resistance only increases the risk of AD through some kind of “transmission” effect, while the peripheral insulin resistance existing in patients without diabetes is not as serious as diabetes, so the “transmission” effect of peripheral insulin resistance is weaker than diabetes.

Insulin resistance can occur before many related diseases. Since studies have confirmed that the degree of CNS is related to a decrease in cognitive function, it can be inferred that CNS occurs earlier than cognitive dysfunction, although the mechanism has not been confirmed. Receptor resistance, which affects certain tissues more than others, is a well-known feature of hormone resistance. Increasing insulin has been shown to improve cognition in AD[29], which may be caused by a lack of insulin action in the CNS, which in turn may be caused by insufficient levels of insulin in the CNS, as well as resistance to insulin at the receptor. As CNS receptors do not partake in the conventional negative feedback mechanism concerning glucose and insulin levels, a deficiency in insulin activity within the CNS has minimal effect on blood insulin levels.

Despite the aforementioned advantages and potential clinical significance, this study still has some limitations that should be taken into account when interpreting the results. Firstly, this is a post hoc analysis, and the initial study was not conducted to examine the relationship between the TyG index and cognitive impairment. Second, the study had a short follow-up time, and this limited the application of the results of this study. In future studies, we hope to explore whether there is an important difference between the TyG index in various cognitive fields.

## CONCLUSION

In this post-hoc analysis using data from the SPRINT, we found that the TyG index was associated with probable dementia in the hypertensive patients, and there was no gender difference between the TyG index and probable dementia. More long-term follow-up studies will be needed to clarify this association. The diagnosis and treatment of TyG index in metabolic or non-metabolic diseases still need further exploration.

## Data Availability

Availability of data and materials Publicly available datasets were analyzed in this study. This data can be found here: https://biolincc.nhlbi.nih.gov/studies/sprint/.

## DECLARATIONS

### Ethics approval and consent to participate

Not applicable

### Consent for publication

Not applicable

### Availability of data and materials

Publicly available datasets were analyzed in this study. This data can be found here: https://biolincc.nhlbi.nih.gov/studies/sprint/.

### Authors’ contributions

RL wrote the paper. RL applied for the database and made statistical analysis. WC was responsible for the revision of the paper. Both authors contributed to the article and approved the submitted version.

## Acknowledgements

We thank Keyang Zheng from the Capital Medical University for assistance in developing the content of this article.

